# Repeatability and reproducibility of in-vivo brain temperature measurements

**DOI:** 10.1101/2020.10.27.20220715

**Authors:** Ayushe A. Sharma, Rodolphe Nenert, Christina Mueller, Andrew Maudsley, Jarred Younger, Jerzy P. Szaflarski

**Author notes:** **Correspondence:** Ayushe Sharma, UAB Epilepsy Center, University of Alabama at Birmingham, Department of Neurology, 1719 6^th^Avenue South, CIRC 312, Birmingham, AL 35249-0021, USA.

## Abstract

**Background:** Magnetic resonance spectroscopic imaging (MRSI) is a neuroimaging technique that can be used to noninvasively map brain temperature (i.e., thermometry) over a large brain volume. To date, intra-subject reproducibility of MRSI-based brain temperature (MRSI-t) has not been investigated. The objective of this repeated measures MRSI-t study was to establish intra-subject reproducibility and repeatability of brain temperature, as well as typical brain temperature range.

**Methods:** Healthy participants aged 23-46 years (N=18; 7 females) were scanned at two time points, ∼12-weeks apart. Volumetric MRSI data were processed by reconstructing metabolite and water images using parametric spectral analysis. Brain temperature was derived using the frequency difference between water and creatine (T_CRE_) for 47 regions of interest (ROIs) delineated by the modified Automated Anatomical Labeling (AAL) atlas. Reproducibility was measured using the coefficient of variation for repeated measures (COVrep), and repeatability was determined using the standard error of measurement (SEM). For each region, the upper and lower bounds of Minimal Detectable Change (MDC) were established to characterize the typical range of T_CRE_ values.

**Results:** The mean global brain temperature over all subjects was 37.2°C, with spatial variations across ROIs. There was a significant main effect for time (*F*(1, 1591)=37.0, *p* < 0.0001) and for brain region (*F*(46, 1591)=2.66, *p*<0.0001). The time*brain region interaction was not significant (*F*(46, 1591)=0.80, *p*=0.83)). Participants’ T_CRE_ was stable for each ROI across both time points, with ROIs’ COVrep ranging from 0.81 – 3.08% (mean COVrep = 1.92%); 30 ROIs had a COVrep < 2.0%.

**Conclusions:** Brain temperature demonstrated subtle regional variations that were highly consistent between both time points, indicating high reproducibility and repeatability of MRSI-t. MRSI-t may be a promising diagnostic, prognostic, and therapeutic tool for non-invasively monitoring pathological brain temperature changes when other modalities are unrevealing. However, further studies of healthy participants with larger sample size(s) and numerous repeated acquisitions are imperative for establishing a reference range of typical brain T_CRE_, as well as the threshold above which T_CRE_ is likely pathological.

## 1 INTRODUCTION

Neuroinflammation is increasingly implicated in the initiation and maintenance of a number of neurodegenerative diseases, including multiple sclerosis, Alzheimer’s Disease, and epilepsy (Lucas et al., 2006; Amor et al., 2010; Maroso et al., 2010). Structural and functional magnetic resonance imaging (MRI) studies are informative but frequently provide an incomplete clinical characterization of focal tissue abnormalities. In the context of surgical intervention for epilepsy management, patients experience worse long-term outcomes when MRI proves unremarkable (Alberts et al.; Salmenpera et al., 2007; Giorgio and De Stefano, 2010; Finke, 2018; Sharma and Szaflarski, 2020). Therefore, there is a dire need to identify a sensitive and specific *in vivo* biomarker of neuroinflammatory pathophysiology. Currently available approaches are invasive and/or costly (e.g., lumbar puncture, positron emission tomography [PET]). Further, PET relies on the use of radioisotopes that *may* localize key neuroinflammatory cells, but variable quantitative accuracy, limited bioavailability, and unclear specificity of ligand-target binding are critical gaps that limit its clinical utility (Hamelin et al.; Best et al., 2019; Vivash and OBrien, 2016; Dickstein et al., 2019; Ghadery et al., 2019; Sharma and Szaflarski, 2020). Thus, the question remains: how can we non-invasively localize neuroinflammation in a living human brain?

Since the focal inflammatory response generates focal changes in temperature, brain temperature mapping may be a promising proxy for diagnosing and monitoring the progression of neurological diseases characterized by metabolic and homeostatic disruptions (Ene Mrozek et al.; Wang et al., 2014). Brain temperature correlates well with systemic temperature during healthy states, generally measuring 0.5–1°C higher than core body temperature (Rossi et al., 2001; Wang et al., 2014). During pathological processes such as neuroinflammation, increased metabolic demands overwhelm the brain’s already limited cooling mechanisms and drive brain temperature by 1-2°C higher than core body temperature (Rossi et al., 2001). As neuroinflammatory phenomena such as leukocyte extravasation and accumulation, blood brain barrier permeability, and even cerebral edema increase, so does brain temperature (Dietrich et al., 1996, 1998; Chatzipanteli et al., 2000; Sharma and Hoopes, 2003).

Of the ways we can measure brain temperature, magnetic resonance spectroscopic imaging (MRSI) is the most non-invasive and economical. Brain temperature can be derived from MRSI data by calculating the frequency difference between the temperature-sensitive water peak and one or more metabolite peaks that are temperature-insensitive (Maudsley et al., 2017). MRSI-based brain temperature (MRSI-t) measurement correlates well with the recordings of implanted probes, as indicated by phantom and experimental studies (Cady et al., 1995; Corbett et al., 1995). Brain temperature has already been approximated using MRSI in a number of contexts: healthy adults, ischemic stroke, neonatal encephalopathy, myalgic encephalitis/chronic fatigue syndrome, and rheumatoid arthritis (Maudsley et al., 2010; Mueller et al., 2019, 2020; Zhang et al., 2020). The majority of previous studies have performed MRSI-t measurements only at a single brain location or limited spatial region, but it has recently been demonstrated that the measurement can be obtained using a volumetric echo-planar spectroscopic imaging (EPSI) acquisition to create temperature maps over a large fraction of the brain volume (Maudsley et al., 2006, 2017). Limited data are available on the reproducibility and reliability of MRSI-t using volumetric EPSI. Previous work has established intra-subject reproducibility of serial EPSI acquisitions in a limited sample (N=2) at 5 one-week intervals, as well as that of successive EPSI acquisitions (2 per session) in a larger sample (N=32) (Maudsley et al., 2010; Veenith et al., 2014). However, the reproducibility and repeatability of MRSI-t using volumetric EPSI has only been established in a small sample (N=10), with 3 acquisitions separated by one-week intervals (Zhang et al., 2020). Thus, the available data do not mirror a real-life clinical situation in which participants are typically exposed to a treatment/intervention for 12-16 weeks before a second measurement is collected.

To date, no study has investigated whether intra-subject reproducibility is maintained in a larger sample of healthy control participants with a longer duration between scans. Additionally, all previous studies have only reported the coefficient of variation (COV) and intraclass correlation coefficients as measures of reproducibility and repeatability; these data are useful, but the exact boundary at which a given region’s brain temperature is normal versus above-normal (i.e., areas of focal neuroinflammation or other pathophysiological phenomena) remains uncertain (Maudsley et al., 2010; Zhang et al., 2020). Before MRSI-t can be used as a clinical tool for in vivo assessment of neuroinflammation, we must determine whether MRSI-based brain temperature estimates are reliable and reproducible over typical study periods. In addition to establishing bounds of normal global and region-level brain temperature, it is critical that we determine the bounds at which brain temperature is above-normal in each region. The objective of this study was to establish intra-subject reproducibility and repeatability of brain temperature derivations in healthy participants scanned twice, approximately 12 weeks apart. In addition to establishing typical mean brain temperature across regions, we aimed to define the bounds of typical vs. atypical (i.e., possibly abnormal) brain temperature by calculating region-level measures of Minimum Detectable Change (MDC). We hypothesized that mean brain temperature would range from 37.0 to 37.5°C. We also hypothesized that MDC would indicate brain temperatures to be atypical if ≥ 0.5°C above mean brain temperature.

## 2 METHODS

### 2.1 Participants

Healthy adult participants were recruited from the local area via word of mouth and IRB-approved study flyers. Interested participants contacted study personnel via phone or email. Participants were scheduled for their first study visit pending a phone screen for inclusion criteria and MR compatibility. The primary inclusionary criteria were: 1) age of 18 to 65 years, 2) ability to undergo MRI at 3-Tesla (e.g., no metal implants or claustrophobia), and 3) negative urine pregnancy test if female of childbearing potential. Written informed consent was obtained from all participants before collecting any clinical measures or imaging data.

### 2.2 Study Visits

Participants completed two study visits scheduled at the University of Alabama at Birmingham (UAB) Civitan International Neuroimaging Laboratory housed in the UAB Highlands Hospital. For most participants (N=14), visits were scheduled 10-12 weeks apart; for the last 4 participants recruited, the second study visit was delayed by 5 - 7 weeks due to COVID-19 research restrictions. The mean overall time between visits was 11.33 ± 2.59 weeks. During each study visit, participants completed 2 self-report questionnaires: the Hospital Anxiety and Depression Scale (HADS) and the Profile of Mood States (POMS) (Zigmond and Snaith, 1983; McNair et al., 1989). Prior to imaging, tympanic temperature measurements were collected using a Braun Pro 4000 ThermoScan aural thermometer.

### 2.3 Data Acquisition

Participants were scanned on a 3T Siemens Magnetom Prisma scanner using a 20-channel head coil. T1-weighted structural images were acquired using a magnetization-prepared rapid gradient echo sequence with the following parameters: repetition time (TR)=2400 ms; echo time (TE)=2.22 ms; flip angle=8°; 208 slices (0.8 mm thick); matrix=256×256. Whole-brain metabolite MRSI data were collected using a 3-dimensional EPSI sequence (TR_1_=1500 ms, TR_2_=511 ms, TE=17.6 ms, lipid inversion-nulling with TI=198 ms; FOV=280×280×180mm; 5.6×5.6×14.4mm voxels). A separate water MRSI dataset was acquired using an interleaved acquisition with identical spatial encoding. We also acquired 2D arterial spin labeling (ASL) perfusion scans to rule out perfusion-related contributions to brain temperature and metabolite concentration measurements (Ene Mrozek et al.; Zhu et al., 2009; Rango et al., 2015). ASL data were acquired using a Proximal Inversion with Control of Off-Resonance Effects (PICORE) labeling scheme for background suppression. Sixty pairs of label/control ASL images were collected in the axial direction at a single inversion time of 1800ms, TR=2500ms, TE=16.18ms, 12 slices, 4×4×10 mm voxels.

### 2.4 Data Processing

Image reconstruction and spectral processing was completed within the Metabolite Imaging and Data Analysis System (MIDAS) software package (Maudsley et al., 2006). Raw metabolite and water MRSI data were reconstructed with interpolation to 64×64×32 and spatial smoothing, resulting in a voxel volume of approximately 1.5 cm^2^. Processing included B_0_ inhomogeneity correction and formation of individual metabolite maps using a parametric spectral modelling method to quantify relative peak areas and resonance frequencies for resonances of N-acetylaspartate (NAA), creatine (CRE), choline, and water, as described by Maudsley et al. (Maudsley et al., 2006). For repeat studies, each MRSI dataset was spatially registered to the T1-weighted image of the first study.

Maps of brain temperature were then calculated using the chemical shift difference between the CRE and water resonances (Δ_H20-CRE□_) according to the equation T_CRE_ =□ −102.61(Δ_H20-CRE□_) + 206.1°C (Maudsley et al., 2017). The CRE resonance was selected for the frequency reference as it is broadly distributed within the cells and as such is less sensitive to cellular-level changes of magnetic susceptibility with neuronal orientation (Maudsley et al., 2017). The resultant MRSI-t maps were then spatially registered to Montreal Neurological Institute (MNI) template at 2mm isotropic voxel resolution, which was also aligned with a modified version of the Automated Anatomical Labeling (AAL) atlas that delineated 47 regions of interest (ROIs) (Tzourio-Mazoyer et al., 2002; Maudsley et al., 2017).

Following initial processing, the atlas was mapped into subject space using an inverse spatial transformation algorithm within the MIDAS Project Review and Analysis (PRANA) module (Maudsley et al., 2006). The Map Integrated Spectrum (MINT) module within MIDAS was then used to compute mean estimates of metabolite concentrations, metabolite ratios, and brain temperature within each of the atlas-defined brain regions. Spectral integration was limited to voxels that had a fitted metabolite linewidth >12 Hz or <2Hz, and voxels were excluded if they had an outlying values of 2.5 times the standard deviation of all valid voxels in the image (Maudsley et al., 2006).

ASL data were processed using ASLtbx batch scripts with Statistical Parametric Mapping (SPM12; http://www.fil.ion.ucl.ac.uk) running in MatLab R2019b (The MathWorks, Inc., Natick, MA, USA) (Wang et al., 2008). Images were motion-corrected and smoothed with a 6 mm full-width-at-half-maximum (FWHM) Gaussian kernel to diminish motion artifacts and decrease noise for subsequent image subtraction. Cerebral blood flow (CBF) was quantified in ml/100g/min using simple subtraction between each tag/control pair (120 smoothed volumes, 60 pairs). Each participant’s mean CBF maps were then 1) registered to high-resolution structural space with affine registration, followed by 2) non-linear registration to MNI space.^i^

### 2.5 Data Analysis

Descriptive statistics and correlation analyses were conducted in IBM SPSS Version 26.0 (www.ibm.com/products/spss-statistics). Reproducibility of MRSI-t based T_CRE_ was evaluated using the coefficient of variation for repeated measures (COVrep) (Shechtman, 2013). Reliability was evaluated with the Standard Error of Measurement (SEM = square root of the MS_ERROR_ term from 2-way mixed ANOVA used to compute Cronbach’s Alpha) (Weir, 2005). Minimal Detectable Change (MDC) was then calculated from SEM to provide a clinically meaningful basis for evaluating T_CRE_ changes over repeated measures. Standard MDC was calculated at the 68^th^ confidence interval (CI) (Weir, 2005). To investigate temporal and spatial variation in T_CRE_, we performed a linear mixed effects analysis using GraphPad Prism version 8.0 for Mac (GraphPad Software, La Jolla, CA, USA, www.graphpad.com).

In a secondary analysis, the relationship between T_CRE_ and metabolites and/or metabolite ratios implicated in neuroinflammatory disease was assessed. Metabolites of interest included myo-inositol, choline, NAA, and the combined signal of glutamine and glutamate (Glx) (Oz et al., 2014). Myo-inositol (MI) is a glial marker, with increased levels indicating glial activation or proliferation seen in neuroinflammation (Haris et al., 2011). Choline (total choline, tCho) is expressed in cell membranes, with increased levels indicating high cell turnover during inflammatory processes (Oz et al., 2014). NAA indicates neuronal health, with lower values representing axonal loss (Moffett et al., 2007; Oz et al., 2014). Due to evidence for creatine (total creatine, tCre) as a reference metabolite, metabolites were evaluated on the basis of their ratios with tCre (Maudsley et al., 2017). Spearman’s rho (*r*_s_) correlation coefficients were calculated between T_CRE_, each metabolite ratio of interest, and tympanic temperature using a two-tailed threshold of p<0.05. Variables with *r*_s_ >|0.5| were evaluated as predictors of T_CRE_ in a multiple regression model. Independent samples t-tests assessed hemispheric (right vs left) differences in region-level T_CRE_. Independent samples t-tests contrasting sex differences in global and region-level T_CRE_ were planned but not performed due to unequal sex distribution in the final dataset. Data quality was evaluated on the basis of 1) number of accepted voxels (%) following processing and 2) spectral linewidth.

Paired samples t-tests of participants’ mean CBF maps contrasted cerebral perfusion between the two time points. This served as a measure of whether brain temperature differences – if present – could be attributed to perfusion differences.

## 3 RESULTS

### 3.1 Participant Demographics and Metabolite Measures

Twenty-one participants were recruited; 18 completed all study procedures and were included in the analyses (N=7 females). The mean age was 30.39 ±7.47 years (range 23–46 years). Descriptive statistics for study measures of temperature, blood flow, and inflammatory metabolite ratios were tabulated for both time points; global within-subject differences from time1 to time2 were computed with repeated measures t-tests (Table 1). The repeated measures t-tests revealed significant increase in HADS sub-scale Depression scores [*t*(17)=-4.12, *p*=0.001], though this change was not clinically significant and scores remained in the normal range (0-7) for both time points (Zigmond and Snaith, 1983). Additionally, there was a global reduction in mean NAA/tCRE, *t*(17)=2.19, *p*=0.04. Mean T_CRE_ was 37.00°C at time1 and 37.40°C at time2, with a global mean T_CRE_ of 37.2°C across both time points. Box-and-whisker plots of T_CRE_ calculated at both time points are provided in Figure 1.

**Table 1.** Descriptive statistics for clinical and imaging-derived measures of temperature, blood flow, or inflammatory processes.

|  | Time 1 (N=18) | Time 2 (N=18) | Time 1 vs Time 2 |
| --- | --- | --- | --- |
| Heart Rate (bpm) | 73.50 ±11.40 | 74.17 ±17.12 | $t(17)=-0.20, p=0.84$ |
| HADS, Depression | 1.72 ±1.96 | 2.72 ±2.27 | $t(17)=-4.12, p=0.001$ |
| HADS, Anxiety | 5.33 ±2.63 | 4.83 ±2.36 | $t(17)=0.89, p=0.39$ |
| POMS, TMD | -0.39 ±20.75 | 0.06 ±18.89 | $t(17)=-0.18, p=0.85$ |
| Tympanic Temperature (°C) | 36.73 ±0.30 | 36.65 ±0.19 | $t(17)=0.98, p=0.34$ |
| Global T<sub>CRE</sub> (°C) | 37.00 ±0.62 | 37.40 ±0.94 | $t(17)=-1.28, p=0.22$ |
| Global CBF (ml/100g/min) | 70.77 ±6.15 | 71.03 ±4.24 | $t(17)=0.21, p=0.84$ |
| tCre | 26270.0 ±1807 | 26844.98<br>±1345.20 | $t(17)=-1.91, p=0.07$ |
| tCho | 5279.00 ±664.10 | 5288.66 ±723.35 | $t(17)=-0.10, p=0.93$ |
| NAA/tCre | 1.35 ±0.24 | 1.26 ±0.09 | $t(17)=2.19, p=0.04$ |
| Glx/tCre | 0.66 ±0.06 | 0.64 ±0.05 | $t(17)=1.76, p=0.10$ |
| MI/tCre | 0.55 ±0.08 | 0.59 ±0.22 | $t(17)=-0.63, p=0.53$ |

\*All measures are in I.U., institutional units unless otherwise specified.
Abbreviations: HADS – Hospital Anxiety and Depression Scale; POMS – Profile of Mood States; TMD – total mood disturbance; CBF, cerebral blood flow; tCHO, total choline; tCre, creatine + phosphocreatine (total creatine); NAA/tCre, N-acetylaspartate + N- acetylaspartyl glutamate ratio to tCre; Glx/tCr, ratio of combined signal from glutamine and glutamate to tCre; MI/tCre, myo-inositol ratio to tCre.

**Figure 1.**
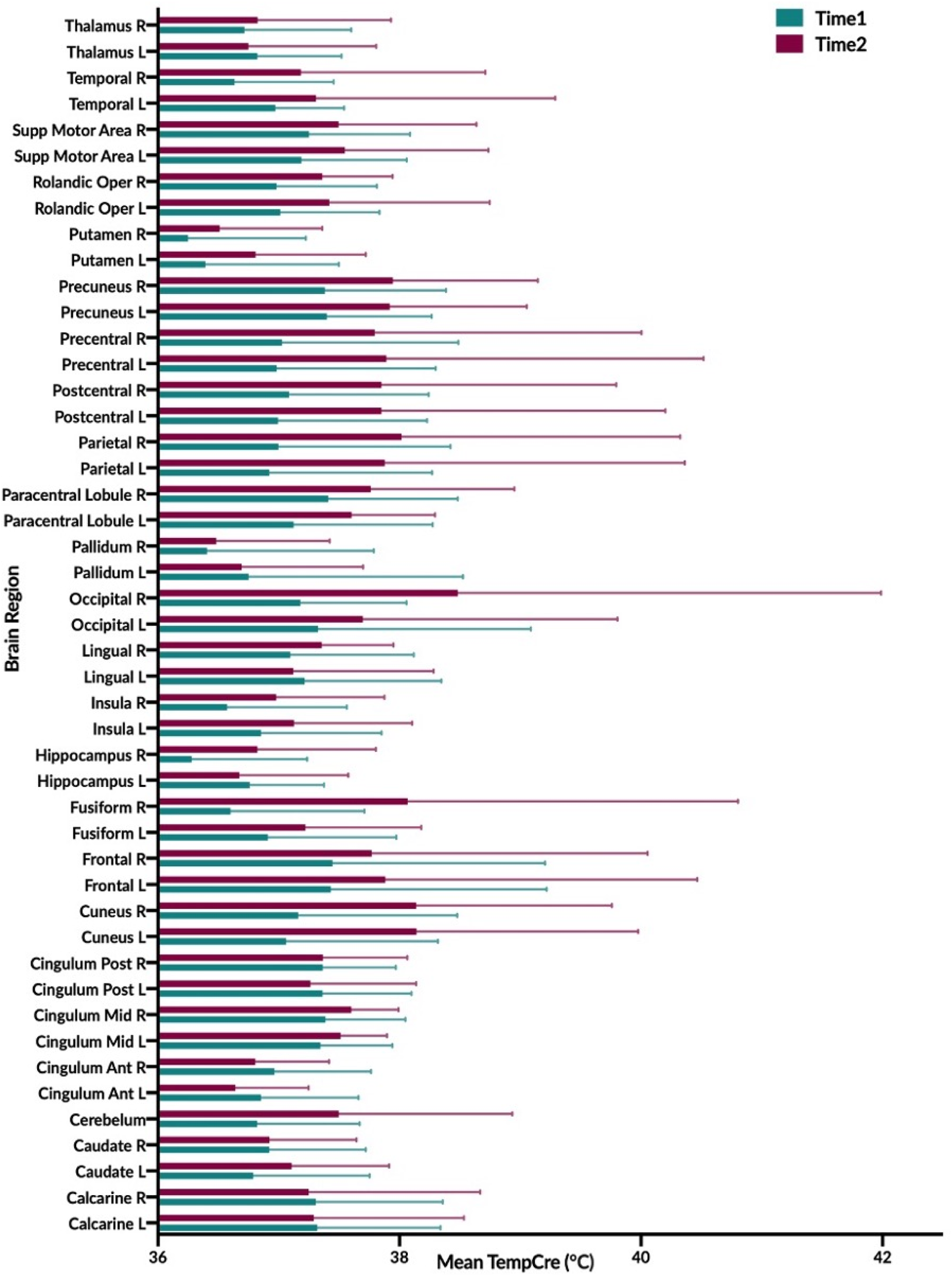
Graphical illustration of mean brain temperature for time1 (mean 37.0°C) and time2 (37.4°C) across 47 regions of interest (ROIs) delineated by the modified Automated Anatomical Labeling (AAL) atlas. Brain temperature was derived using the frequency difference between water and creatine (T_CRE_) following processing of volumetric MRSI data. For each brain region, colored bars (Time1, teal; Time2, burgundy), reflect the mean T_CRE_; whiskers indicate range of T_CRE_ values beyond the mean. The mean global brain temperature over all subjects was 37.2°C with spatial variations across ROIs (37.57 - 41.49°C). There was a significant main effect for time (*F*(1, 1591)=37.0, *p* < 0.0001) and for brain region (*F*(46, 1591)=2.66, *p*<0.0001). The time*brain region interaction was not significant (*F*(46, 1591)=0.80, *p*=0.83)). Abbreviations: MRSI, magnetic resonance spectroscopic imaging; ROI, region of interest; n, number of observations; R, right; L, left.

### 3.2 Brain Temperature Reproducibility, Reliability, and Minimal Detectable Change

The COVrep, our measure of reproducibility, ranged from 0.81 – 3.08% (mean 1.92%) across 47 ROIs, with 30 ROIs having a COVrep < 2.0%. SEM ranged from 0.365 – 2.713 (mean 1.295 ± 0.647) (Table 2). The highest COVrep (3.08%) was in the R Fusiform Gyrus. Mean brain temperature was the lowest in the R and L putamen, R and L pallidum, R and L hippocampus, L and R anterior cingulum, R and L thalamus, and R insula (Table 2). Mean brain temperature was highest in the R occipital lobe, L and R precuneus, R and L frontal lobes, and R and L cuneus.

**Table 2.** Mean MRSI-based brain temperature (T_CRE_) in 47 ROIs for both time points time1 (T_1_) and time2 (T_2_), their standard deviations (SD), and corresponding reproducibility and reliability statistics. For each ROI, reproducibility was measured with the coefficient of variation for repeated measures (COVrep). The standard error of measurement (SEM) provided a measure of reliability. Minimal Detectable Change (MDC) was calculated from SEM to provide a clinically meaningful basis for evaluating T_CRE_ changes over repeated measures.

| ROI | Scan | n | Mean | SD | COVrep (%) | SEM | MDC |
| --- | --- | --- | --- | --- | --- | --- | --- |
| **L Calcarine** | T<sub>1</sub> | 18 | 37.320 | 1.019 | 1.85 | 1.131 | 1.600 |
|  | T<sub>2</sub> | 18 | 37.289 | 1.244 |  |  |  |
| **R Calcarine** | T<sub>1</sub> | 18 | 37.306 | 1.052 | 2.24 | 1.288 | 1.822 |
|  | T<sub>2</sub> | 18 | 37.247 | 1.421 |  |  |  |
| **L Caudate** | T<sub>1</sub> | 18 | 36.788 | 0.963 | 1.66 | 0.733 | 1.036 |
|  | T<sub>2</sub> | 18 | 37.106 | 0.807 |  |  |  |
| **R Caudate** | T<sub>1</sub> | 18 | 36.921 | 0.800 | 1.73 | 0.827 | 1.170 |
|  | T<sub>2</sub> | 18 | 36.922 | 0.721 |  |  |  |
| **Cerebellum** | T<sub>1</sub> | 18 | 36.822 | 0.848 | 2.10 | 1.141 | 1.614 |
|  | T<sub>2</sub> | 18 | 37.497 | 1.436 |  |  |  |
| **L Anterior Cingulum** | T<sub>1</sub> | 17 | 36.853 | 0.807 | 1.29 | 0.581 | 0.822 |
|  | T<sub>2</sub> | 17 | 36.641 | 0.606 |  |  |  |
| **R Anterior Cingulum** | T<sub>1</sub> | 17 | 36.965 | 0.799 | 1.11 | 0.511 | 0.722 |
|  | T<sub>2</sub> | 17 | 36.806 | 0.610 |  |  |  |
| **L Mid Cingulum** | T<sub>1</sub> | 18 | 37.345 | 0.597 | 0.81 | 0.365 | 0.516 |
|  | T<sub>2</sub> | 18 | 37.512 | 0.383 |  |  |  |
| **R Mid Cingulum** | T<sub>1</sub> | 18 | 37.387 | 0.661 | 0.88 | 0.421 | 0.595 |
|  | T<sub>2</sub> | 18 | 37.600 | 0.391 |  |  |  |
| **L Posterior Cingulum** | T<sub>1</sub> | 18 | 37.363 | 0.735 | 1.41 | 0.824 | 1.165 |
|  | T<sub>2</sub> | 18 | 37.262 | 0.875 |  |  |  |
| **R Posterior Cingulum** | T<sub>1</sub> | 18 | 37.364 | 0.605 | 1.36 | 0.696 | 0.985 |
|  | T<sub>2</sub> | 18 | 37.366 | 0.698 |  |  |  |
| **L Cuneus** | T<sub>1</sub> | 18 | 37.062 | 1.257 | 2.50 | 1.522 | 2.153 |
|  | T<sub>2</sub> | 18 | 38.140 | 1.835 |  |  |  |
| **R Cuneus** | T<sub>1</sub> | 18 | 37.161 | 1.316 | 2.65 | 1.586 | 2.243 |
|  | T<sub>2</sub> | 18 | 38.138 | 1.620 |  |  |  |
| **L Frontal Lobe** | T<sub>1</sub> | 18 | 37.431 | 1.787 | 2.88 | 2.713 | 3.837 |
|  | T<sub>2</sub> | 18 | 37.884 | 2.582 |  |  |  |
| **R Frontal Lobe** | T<sub>1</sub> | 17 | 37.445 | 1.759 | 2.56 | 2.395 | 3.387 |
|  | T<sub>2</sub> | 17 | 37.771 | 2.285 |  |  |  |
| **L Fusiform Gyrus** | T<sub>1</sub> | 18 | 36.909 | 1.065 | 1.60 | 0.831 | 1.176 |
|  | T<sub>2</sub> | 18 | 37.221 | 0.959 |  |  |  |
| **R Fusiform Gyrus** | T<sub>1</sub> | 18 | 36.601 | 1.108 | 3.08 | 2.179 | 3.082 |
|  | T<sub>2</sub> | 18 | 38.067 | 2.735 |  |  |  |
| **L Hippocampus** | T<sub>1</sub> | 18 | 36.758 | 0.616 | 1.55 | 0.879 | 1.243 |
|  | T<sub>2</sub> | 18 | 36.674 | 0.902 |  |  |  |
| **R Hippocampus** | T<sub>1</sub> | 18 | 36.278 | 0.956 | 2.13 | 1.016 | 1.437 |
|  | T<sub>2</sub> | 18 | 36.824 | 0.980 |  |  |  |
| **L Insula** | T<sub>1</sub> | 18 | 36.854 | 0.996 | 1.92 | 1.022 | 1.446 |
|  | T<sub>2</sub> | 18 | 37.126 | 0.978 |  |  |  |
| **R Insula** | T<sub>1</sub> | 18 | 36.572 | 0.991 | 1.93 | 0.934 | 1.321 |
|  | T<sub>2</sub> | 18 | 36.979 | 0.897 |  |  |  |
| **L Lingual Gyrus** | T<sub>1</sub> | 18 | 37.213 | 1.134 | 1.89 | 1.111 | 1.572 |
|  | T<sub>2</sub> | 18 | 37.120 | 1.161 |  |  |  |
| **R Lingual Gyrus** | T<sub>1</sub> | 18 | 37.096 | 1.021 | 1.61 | 0.853 | 1.206 |
|  | T<sub>2</sub> | 18 | 37.355 | 0.594 |  |  |  |
| **L Occipital Lobe** | T<sub>1</sub> | 18 | 37.326 | 1.761 | 1.33 | 0.731 | 1.033 |
|  | T<sub>2</sub> | 18 | 37.696 | 2.106 |  |  |  |
| **R Occipital Lobe** | T<sub>1</sub> | 18 | 37.179 | 0.879 | 2.98 | 2.507 | 3.546 |
|  | T<sub>2</sub> | 18 | 38.484 | 3.504 |  |  |  |
| **L Pallidum** | T<sub>1</sub> | 18 | 36.749 | 1.776 | 2.58 | 1.195 | 1.691 |
|  | T<sub>2</sub> | 18 | 36.693 | 1.005 |  |  |  |
| **R Pallidum** | T<sub>1</sub> | 17 | 36.406 | 1.381 | 2.70 | 1.292 | 1.828 |
|  | T<sub>2</sub> | 17 | 36.483 | 0.939 |  |  |  |
| **L Paracentral Lobule** | T<sub>1</sub> | 18 | 37.124 | 1.151 | 1.49 | 0.663 | 0.937 |
|  | T<sub>2</sub> | 18 | 37.603 | 0.692 |  |  |  |
| **R Paracentral Lobule** | T<sub>1</sub> | 18 | 37.409 | 1.073 | 1.66 | 1.430 | 2.023 |
|  | T<sub>2</sub> | 18 | 37.761 | 1.190 |  |  |  |
| **L Parietal Lobe** | T<sub>1</sub> | 18 | 36.921 | 1.349 | 2.57 | 2.564 | 3.627 |
|  | T<sub>2</sub> | 18 | 37.877 | 2.485 |  |  |  |
| **R Parietal Lobe** | T<sub>1</sub> | 18 | 36.998 | 1.424 | 2.47 | 2.414 | 3.414 |
|  | T<sub>2</sub> | 18 | 38.016 | 2.307 |  |  |  |
| **L Postcentral Gyrus** | T<sub>1</sub> | 18 | 36.994 | 1.232 | 2.35 | 2.420 | 3.422 |
|  | T<sub>2</sub> | 18 | 37.851 | 2.350 |  |  |  |
| **R Postcentral Gyrus** | T<sub>1</sub> | 18 | 37.084 | 1.157 | 1.88 | 2.003 | 2.833 |
|  | T<sub>2</sub> | 18 | 37.850 | 1.944 |  |  |  |
| **L Precentral Gyrus** | T<sub>1</sub> | 18 | 36.984 | 1.315 | 2.57 | 2.609 | 3.690 |
|  | T<sub>2</sub> | 18 | 37.891 | 2.625 |  |  |  |
| **R Precentral Gyrus** | T<sub>1</sub> | 18 | 37.028 | 1.459 | 2.40 | 2.216 | 3.133 |
|  | T<sub>2</sub> | 18 | 37.793 | 2.211 |  |  |  |
| **L Precuneus** | T<sub>1</sub> | 18 | 37.399 | 0.867 | 1.69 | 1.222 | 1.728 |
|  | T<sub>2</sub> | 18 | 37.920 | 1.134 |  |  |  |
| **R Precuneus** | T<sub>1</sub> | 18 | 37.382 | 1.002 | 1.73 | 1.341 | 1.897 |
|  | T<sub>2</sub> | 18 | 37.944 | 1.201 |  |  |  |
| **L Putamen** | T<sub>1</sub> | 18 | 36.392 | 1.105 | 2.18 | 1.029 | 1.455 |
|  | T<sub>2</sub> | 18 | 36.808 | 0.911 |  |  |  |
| **R Putamen** | T<sub>1</sub> | 18 | 36.248 | 0.975 | 1.89 | 0.872 | 1.233 |
|  | T<sub>2</sub> | 18 | 36.511 | 0.849 |  |  |  |
| **L Rolandic Operculum** | T<sub>1</sub> | 18 | 37.013 | 0.821 | 1.76 | 1.289 | 1.823 |
|  | T<sub>2</sub> | 18 | 37.419 | 1.328 |  |  |  |
| **R Rolandic Operculum** | T<sub>1</sub> | 18 | 36.983 | 0.830 | 1.51 | 0.757 | 1.071 |
|  | T<sub>2</sub> | 18 | 37.360 | 0.582 |  |  |  |
| **L Supplemental Motor Area** | T<sub>1</sub> | 18 | 37.187 | 0.872 | 1.66 | 1.301 | 1.840 |
|  | T<sub>2</sub> | 18 | 37.547 | 1.190 |  |  |  |
| **R Supplemental Motor Area** | T<sub>1</sub> | 17 | 37.251 | 0.837 | 1.67 | 1.295 | 1.831 |
|  | T<sub>2</sub> | 17 | 37.496 | 1.142 |  |  |  |
| **L Temporal Lobe** | T<sub>1</sub> | 18 | 36.971 | 0.569 | 1.24 | 1.111 | 1.571 |
|  | T<sub>2</sub> | 18 | 37.308 | 1.981 |  |  |  |
| **R Temporal Lobe** | T<sub>1</sub> | 18 | 36.634 | 0.822 | 1.72 | 1.197 | 1.694 |
|  | T<sub>2</sub> | 18 | 37.182 | 1.529 |  |  |  |
| **L Thalamus** | T<sub>1</sub> | 18 | 36.823 | 0.696 | 1.57 | 0.853 | 1.207 |
|  | T<sub>2</sub> | 18 | 36.748 | 1.059 |  |  |  |
| **R Thalamus** | T<sub>1</sub> | 18 | 36.715 | 0.886 | 1.94 | 1.004 | 1.420 |
|  | T<sub>2</sub> | 18 | 36.824 | 1.104 |  |  |  |

Abbreviations: MRSI, magnetic resonance spectroscopic imaging; ROI, region of interest; n, number of observations; R, right; L, left.

Based on the MDC, the T_CRE_ at which we consider brain temperature as atypical varies across brain regions (Table 2, Figure 2). When considering MDC computations at the 68^th^ CI, the upper bounds of brain temperature indicating above-typical T_CRE_ ranged from 37.57 - 41.49°C (mean 39.03 ± 1.14 °C); the lower bounds indicating below-typical T_CRE_ ranged from 33.74 – 36.91°C (mean 35.37 ± 0.80 °C). According to MDC calculations, the T_CRE_ at which values are considered above-typical were highest in the following regions: R and L frontal lobes (41.49°C, 40.99°C), R occipital lobe (41.38°C), L precentral gyrus (41.13°C), and L and R parietal lobes (40.92°C, 41.03°C) (Table 2, Figure 2). In addition to assessing these ROI-based measures, brain temperature maps visualized brain temperature changes within participants from time1 to time2 (Figure 3).

**Figure 2.**
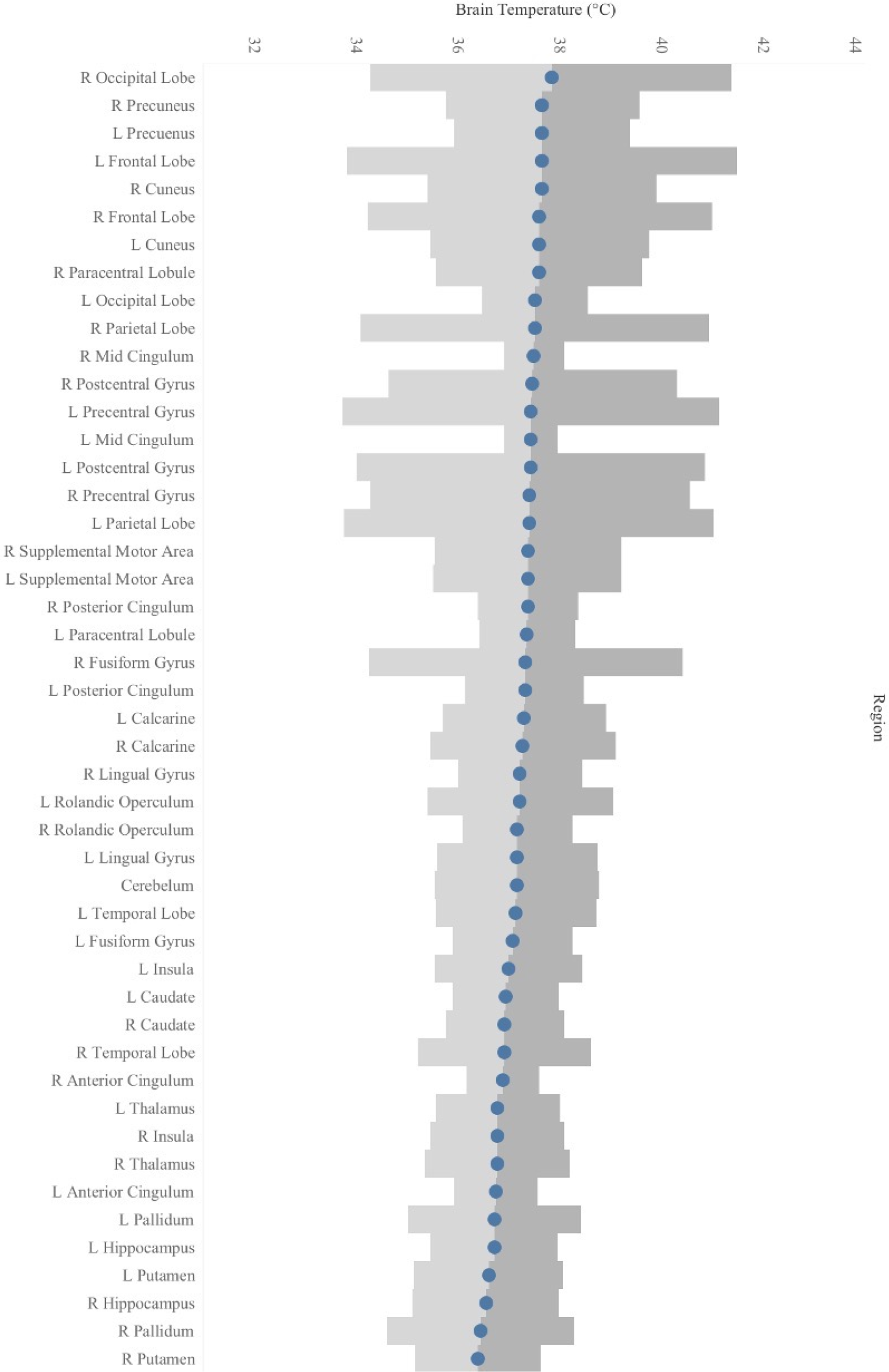
Mean T_CRE_ for each ROI, with Minimal Detectable Change (MDC) upper bounds indicated in dark gray bands and MDC lower bounds indicated in light gray bands.

**Figure 3.**
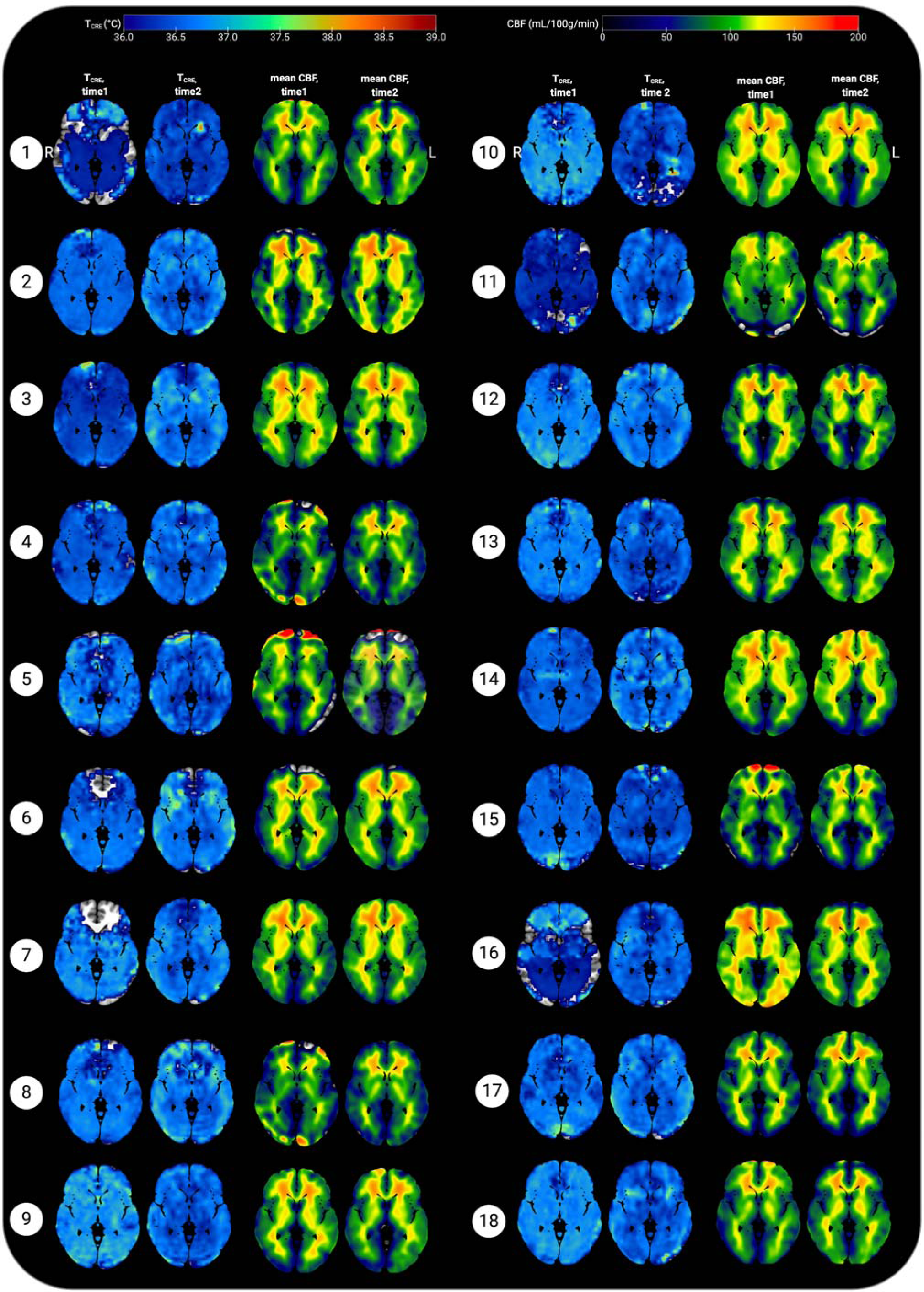
Representative axial slices showing brain temperature (T_CRE)_ and cerebral blood flow (CBF) for 18 healthy participants at time1 and time 2. T_CRE_ and mean CBF maps were resliced and co-registered to MNI space in SPM12 running in MatLab R2017b. The open-source software MRIcroGL (McCausland Center for Brain Imaging, University of South Carolina; https://www.mccauslandcenter.sc.edu/mricrogl/) was used to overlay mean T_CRE_ and CBF maps on the MNI single-participant template for 3D renderings of participant data. The figure was created using BioRender. All data are visualized for axial view of the same slice (z = 8; L, left; R; right). The T_CRE_ color scale ranges from 36.0 – 39°C, with dark to light blue coloring indicating T_CRE_ values within the typical range. The majority of voxels demonstrate T_CRE_ <37.5°C, with a mean of 37.0°C for time1 and 37.4°C for time2 (global mean of 37.2°C). As demonstrated by the time1 and time2 maps and the global COVrep of 1.92%, there was a minimal variation in participants’ data from time1 to time2. For some participants’ data, brain regions were excluded from analysis due to technical reasons; these areas are represented by regions where the template brain is exposed. Each participant’s mean CBF maps for time1 and time 2 are visualized alongside T_CRE_ maps; as with T_CRE_ mean CBF varies little from time1 to time2. Spatial T_CRE_ variations did not vary as a function of cerebral perfusion as indicated by mean CBF.

### 3.4 Spatial and Temporal Variations in Brain Temperature

A linear mixed effects model investigated the variation of T_CRE_ across 47 brain regions over 2 time points. Fixed effects included time, brain region, and the interaction of time*brain region, with participants considered random effects. In the case of a statistically significant interaction, we planned follow-up tests to assess pairwise differences using simple effects analysis. P-values were corrected for multiple comparisons by controlling the False Discovery Rate (FDR=0.05) with the two-stage step-up method of Benjamini, Krieger, and Yekutieli (Benjamini and Yekutieli, 2001). The interaction of time*brain region was not significant (*F*(46, 1591)=0.80, *p*=0.83). There was a significant main effect for time (*F*(1, 1591)=37.0, *p*<0.0001), and for brain region (*F*(46, 1591)=2.66, *p*<0.0001).

### 3.4 Within-Subjects Variation in Cerebral Blood Flow

Voxel-level repeated measures t-tests did not find significant within-subjects differences in mean CBF when comparing time1 vs time 2; the same results were found when mean T_CRE_ was included as a covariate. Further, participants’ global CBF values did not change significantly from time1 to time2, as indicated by paired samples t-tests [*t*(17)=0.21, *p*=0.84]. At time1, mean global CBF was 70.77 ±6.15 mL/100g/min. At time2, mean global CBF was 71.03 ±4.24 mL/100g/min (Table 1). Mean CBF ranged from 56.66 – 79.99 mL/100g/min (Figure 3). Each participant’s mean CBF maps for time1 and time2 are visualized alongside T_CRE_ maps (Figure 3). Mean CBF for all participants across both time points was 70.90 mL/100g/min.

### 3.5 Relationship between Brain Temperature and Other Physiological Variables

For major metabolites (tCre, tCho, NAA, and MI), COVrep (%) values are provided in Table 4. In contrast to T_CRE_, metabolite concentrations varied substantially across time. For tCho, mean COVrep was 12.49% (range 3.13 – 84.52%); with the removal of outlying values for the R Precentral Gyrus, the COVrep range for tCho becomes limited to 3.13 – 34.44%. For tCre, mean COVrep was 10.38% (range 4.35-22.08%). For NAA, mean COVrep was 6.93% (range 2.68 – 28.44%). Lastly, mean COVrep for MI was 16.05% (range 6.41-41.97%). The mean distribution of tCre, tCho, and NAA across regions at time1 is included in Figure 4.

**Table 4.** Coefficient of variation (COVrep, %) for major metabolites.

| Brain Region | COVrep (%) |  |  |  |
| --- | --- | --- | --- | --- |
|  | tCho | tCre | MI | NAA |
| L Calcarine | 7.27 | 8.94 | 5.47 | 13.43 |
| R Calcarine | 11.63 | 14.42 | 10.59 | 13.00 |
| L Caudate | 12.03 | 7.93 | 8.53 | 22.89 |
| R Caudate | 19.00 | 10.67 | 6.62 | 24.31 |
| Cerebelum | 10.61 | 17.31 | 28.44 | 20.28 |
| L Anterior Cingulum | 6.20 | 8.78 | 4.71 | 14.59 |
| R Anterior Cingulum | 6.85 | 6.41 | 3.43 | 12.83 |
| L Mid Cingulum | 9.62 | 9.44 | 3.18 | 13.89 |
| R Mid Cingulum | 5.26 | 11.24 | 2.68 | 17.67 |
| L Posterior Cingulum | 4.73 | 6.45 | 5.01 | 7.19 |
| R Posterior Cingulum | 10.62 | 6.10 | 4.58 | 6.41 |
| L Cuneus | 7.29 | 10.71 | 7.58 | 9.37 |
| R Cuneus | 8.04 | 12.06 | 11.11 | 11.07 |
| L Frontal Lobe | 4.33 | 7.18 | 4.76 | 8.68 |
| R Frontal Lobe | 12.79 | 4.39 | 3.28 | 9.58 |
| L Fusiform Gyrus | 15.91 | 10.46 | 10.02 | 18.75 |
| R Fusiform Gyrus | 10.84 | 13.19 | 19.78 | 20.94 |
| L Hippocampus | 12.45 | 11.14 | 7.72 | 11.01 |
| R Hippocampus | 15.24 | 10.46 | 10.74 | 12.08 |
| L Insula | 3.13 | 6.79 | 3.48 | 19.45 |
| R Insula | 34.44 | 6.60 | 4.63 | 17.07 |
| L Lingual Gyrus | 10.53 | 12.34 | 10.68 | 17.38 |
| R Lingual Gyrus | 16.60 | 13.27 | 13.28 | 16.60 |
| L Occipital Lobe | 5.76 | 9.93 | 6.63 | 11.15 |
| R Occipital Lobe | 9.08 | 12.74 | 8.48 | 9.53 |
| L Pallidum | 12.41 | 10.15 | 8.04 | 41.97 |
| R Pallidum | 12.70 | 17.14 | 8.88 | 37.91 |
| L Paracentral Lobule | 17.79 | 17.57 | 6.26 | 22.12 |
| R Paracentral Lobule | 16.46 | 20.08 | 5.93 | 29.10 |
| L Parietal Lobe | 4.47 | 6.30 | 3.49 | 6.54 |
| R Parietal Lobe | 10.22 | 5.44 | 4.29 | 6.77 |
| L Postcentral Gyrus | 10.16 | 10.10 | 4.18 | 9.89 |
| R Postcentral Gyrus | 10.58 | 11.64 | 4.13 | 12.59 |
| L Precentral Gyrus | 11.91 | 12.20 | 4.56 | 16.17 |
| R Precentral Gyrus | 84.52 | 11.54 | 4.72 | 14.53 |
| L Precuneus | 6.94 | 5.99 | 4.21 | 7.87 |
| R Precuneus | 7.82 | 6.65 | 3.09 | 10.72 |
| L Putamen | 6.23 | 6.94 | 4.91 | 30.85 |
| R Putamen | 13.48 | 9.99 | 7.32 | 22.54 |
| L Rolandic Operculum | 4.07 | 6.96 | 4.25 | 7.96 |
| R Rolandic Operculum | 4.72 | 8.29 | 4.30 | 9.05 |
| L Supplemental Motor Area | 14.95 | 19.46 | 7.08 | 32.60 |
| R Supplemental Motor Area | 22.56 | 22.08 | 4.86 | 25.93 |
| L Temporal Lobe | 3.70 | 4.35 | 3.78 | 8.12 |
| R Temporal Lobe | 8.23 | 5.15 | 6.17 | 6.48 |
| **L Thalamus** | 11.69 | 8.28 | 7.63 | 14.71 |
| **R Thalamus** | 21.24 | 12.43 | 8.35 | 20.60 |

Abbreviations: L, left; R, right; tCHO, total choline; tCre, creatine + phosphocreatine (total creatine); MI, myo-inositol; NAA, N-acetylaspartate + N- acetylaspartyl glutamate ratio.

**Figure 4.**
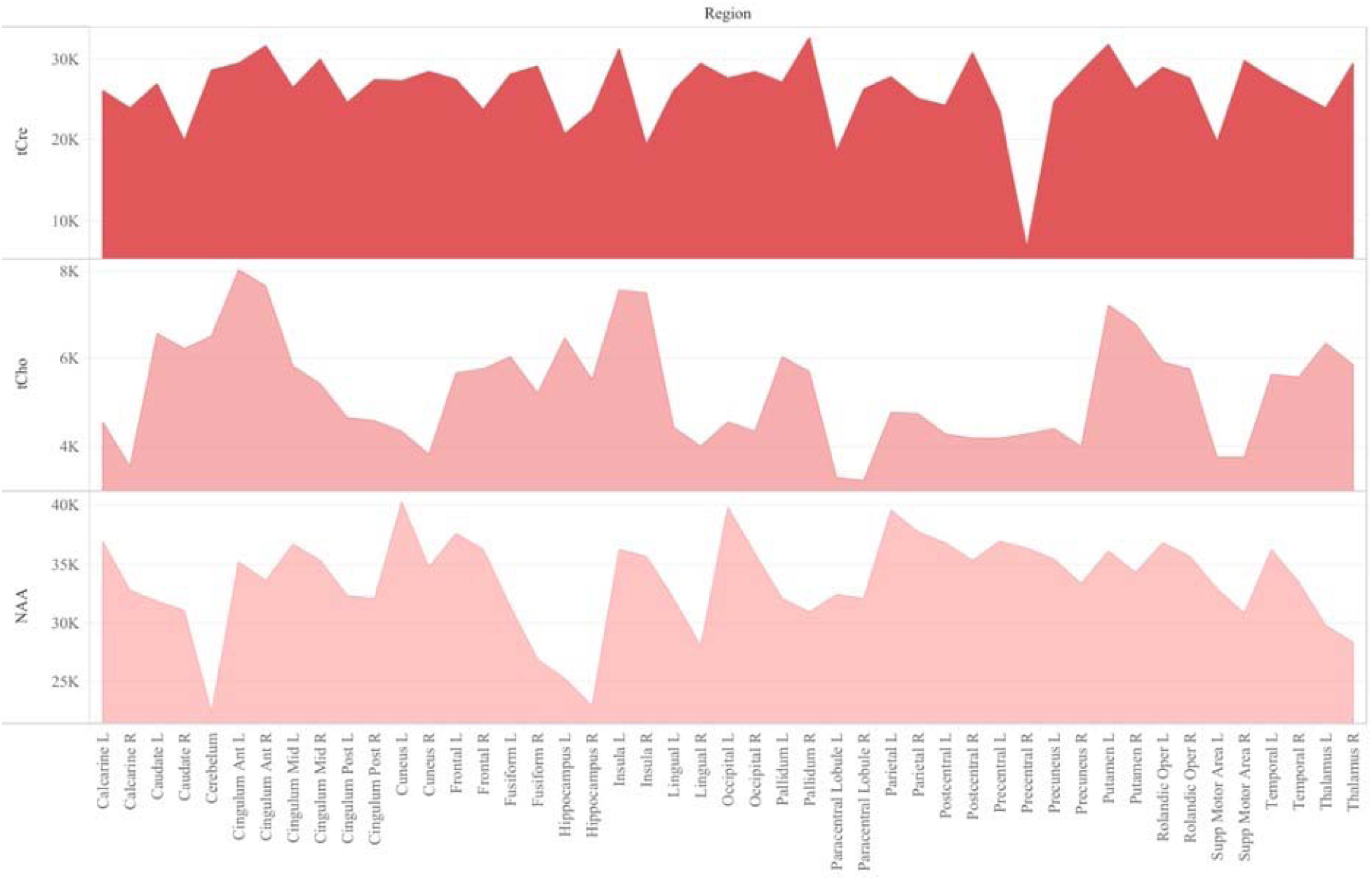
Regional distribution of reference metabolites N-Acetylaspartate (NAA), creatine (total creatine, tCre), and choline (total choline, tCho) at time 1.

Spearman correlations were run to test the relationship between T_CRE_ and the ratios MI/tCRE, NAA/tCRE, and Glx/tCRE (Table 5). There was a strong, positive correlation between T_CRE_ and NAA/tCRE (*r*_s_=0.678, *p*<0.0001), and a strong negative correlation between T_CRE_ and tCho/tCRE (*r*_s_=-0.575, *p*<0.0001). There was a moderate, positive statistically significant correlation between T_CRE_ and Glx/tCRE, *r*_s_=0.458, *p*=0.001. Lastly, there was a weak, negative correlation of statistical significance between T_CRE_ and MI/tCRE, *r*_s_=-0.322, *p*=0.027. Based on results of correlation analysis, a multiple regression was performed to predict T_CRE_ from tCHO/tCRE, NAA/tCRE, Glx/tCRE and MI/tCRE. However, none of the metabolite ratios were significant predictors of T_CRE_.

There were no significant differences in brain temperature when comparing left and right ROIs at p<0.05, corrected for FDR. Due to unequal sex distribution of our participants in the final dataset, we did not perform independent samples t-tests to evaluate sex differences in global and regional T_CRE_.

### 3.6 Assessing Quality and Spectral Resolution

Our data were of moderate to high quality as indicated by the spectral resolution and total percentage of voxels included in our final analysis. An average of 75.18 % of voxels within the brain met quality criteria across both timepoints (76.47 ± 10.68% of voxels at T_1_, 73.89 ± 12.39% of voxels at T_2_), with a range of 48.13% to 89.53%. As indicated by the mean linewidth of 7.31 Hz (range, 6.39 – 8.69), our data also had high spectral resolution.

We performed *post-hoc* correlation analysis of SEM, COVrep, mean T_CRE_, and ROI size to determine if there was an association between reproducibility, repeatability, T_CRE_, and region size. ROI size was calculated by multiplying the mean # of voxels in each region (pre-processing) by the mean % accepted voxels after processing. To adjust for varying scale, SEM, COVrep, mean T_CRE_, and ROI size were z-transformed. Reproducibility and repeatability (COVrep*SEM) had a strong positive correlation (*r*=0.805, *p*<0.001) (Table 6). Mean T_CRE_ had a moderate positive correlation with both SEM (r=0.487, *p*<0.001) and ROI size (r=0.626, *p*<0.001) (Table 6). Finally, in addition to its association with mean T_CRE_, ROI size had a moderate positive correlation with SEM (*r*=0.524, *p*<0.001) (Table 6).

**Table 6.** Pearson correlations between measures of reproducibility and repeatability (SEM and COVrep), mean T_CRE_, and ROI size. All correlations significant at the 0.01 level (2-tailed).

|  | **SEM,**<br>**z-score** | **COVrep,**<br>**z-score** | **Mean T<sub>CRE</sub>,**<br>**z-score** | **ROI Size,**<br>**z-score** |
| --- | --- | --- | --- | --- |
| **SEM, z-score** | - | 0.805 | 0.487 | 0.524 |
| **COVrep, z-score** | 0.805 | - |  |  |
| **Mean T<sub>CRE</sub>, z-score** | 0.487 |  | - | 0.626 |
| **ROI size, z-score** | 0.524 |  | 0.626 | - |

## 4 DISCUSSION

### 4.1 Main Findings

In this study, a global brain temperature of 37.2°C was found, with spatial variations across ROIs consistent with previous studies (Cady et al., 1995, 2011; Zhang et al., 2020). Also consistent with previous studies was a significant main effect for time (*F*(1, 1591)=37.0, *p*<0.0001), and for brain region (*F*(46, 1591)=2.66, *p*<0.0001). The central aim of this study was to evaluate whether intra-subject reproducibility is maintained in a large sample of healthy participants with longer duration between scans than previously investigated. Though this study acquired MRSI-t data with a much greater time interval between acquisitions (∼12 weeks apart), the COVrep ranged from 0.81 – 3.08% (mean COVrep = 1.92%), with 30 ROIs having a COVrep < 2.0%. Thus, T_CRE_ was stable across all 47 ROIs and paralleled results of previous studies of serial acquisitions or those separated by 1-week intervals (Maudsley et al., 2010; Thrippleton et al., 2014; Zhang et al., 2020). Another important question was whether timepoint has an impact on the mean brain temperature within a given ROI. Mixed effects analysis confirmed that brain temperature does change within ROIs as a function of time, as indicated by the lack of a significant interaction (time*brain region, *F*(46, 1591)=0.80, *p*=0.83)). Interestingly, the highest T_CRE_ was found in 1) posterior regions affected by anterior-posterior gradient effects (L and R occipital lobe, L and R cuneus, L and R precuneus), and 2) large peripheral cortical regions (L and R frontal lobes, R parietal, L and R occipital). Large posterior regions demonstrated higher T_CRE_ with more within-subject variability and greater SEM. Based on MDC, atypical T_CRE_ ranged from 37.57 – 41.49°C. MDC was calculated at the 68^th^ confidence interval (CI), but future work with larger sample sizes and more repeated acquisitions may enable MDC calculation at the 90^th^ or 95^th^ CIs. Based on the aforementioned findings, our study confirms previous findings of spatial brain temperature variations from structure to structure; even if time has a significant effect, this effect is distributed over regions, and does not change in magnitude as a function of ROI. Though T_CRE_ correlated moderately with some of the neuroinflammatory metabolites, the regression model indicated that none were significant predictors. Thus, the relationship between T_CRE_ and neuroinflammatory metabolites could not be fully assessed in this population, as would be expected for healthy participants without neuroinflammatory pathophysiology.

Maudsley et al. previously established tCRE as the reference metabolite for brain temperature derivations; this is because of its even distribution across cellular compartments (at least in the absence of disease), which renders tCRE the least susceptible to grey matter (GM) and white matter (WM) tissue-dependent frequency shifts (Maudsley et al., 2017). Given the difficulty of separating GM and WM within each ROI, we used tCRE as our reference metabolite. However, it is not always the case that tCRE is well-distributed or best reference metabolite – especially when considering neurodegenerative conditions characterized by significant changes in bioenergetics/metabolism. The posterior regions that indicated higher brain temperature and greater variability could be the result of anterior-posterior gradient due to acquisition, but may also be the result of visual stimulation effects from watching movies during scanning (Kauppinen et al., 2008; Rango et al., 2015). Future studies would benefit from acquiring data both with and without in-scanner visual stimulation.

### 4.2 Limitations and Other Considerations

Our study was limited by the heterogeneity of the participant population, limited age range (no participants >46 years), and acquisition-related methods that are inherently variable between- and within-participants. Additionally, we did not monitor or control for a number of variables that could alter brain temperature, including circadian rhythms, diurnal changes, hormonal variations (e.g., due to menstruation), or even environmental conditions (e.g., temperature in the scanner room). Our methodological limitations stem from two issues that critically impact all MRSI-t data: 1) magnetic field inhomogeneities and 2) interfering signal from water-containing structures (e.g., the aqueous humor of the eyes). Shimming during EPSI acquisition can substantially reduce magnetic field inhomogeneities and improve signal-to-noise ratio by adjusting spectral linewidth, but the process is time-consuming and difficult without substantial training. Though shimming greatly improves spectral linewidth, it cannot correct the spatial inhomogeneities present across structures and even within large brain regions – additionally, it is unclear whether these spatial inhomogeneities are truly artifact or a reflection of actual physiological processes. Signals from water-containing structures are typically masked with the placement of a saturation band placement during data acquisition. While this is effective to an extent, the saturation band is a 3D rectangular slab of fixed bounds and shape – it can only be angled to cover the eyes and sphenoidal sinuses, and there is currently no capacity for altering the curvature of the band. Depending on each participant’s structural anatomy, the saturation band may cut off regions from regions of cortex in the frontal areas. Additionally, the placement and angling of this slab cannot be replicated from subject-to-subject or even within a subject across time points. As with our own data, these issues can result in missing or even heavily contaminated metabolite and temperature data for impacted ROIs, as voxels may not contain sufficient information or signals of sufficient quality.

A countless number of phenomena could theoretically impact brain temperature and weaken the ability to maintain sufficient reproducibility and reliability. Functional activity, time of day, and even transient hormonal fluctuations such as menstruation may impact MRSI-t-based estimates (Ene Mrozek et al.). Thrippleton et al limited data acquisition to afternoon hours to minimize diurnal temperature variation, and went so far as to recruit only male subjects to avoid the hormonal fluctuations that may impact brain temperature in females (Thrippleton et al., 2014). They additionally instructed their participants to refrain from eating, drinking, exercising, or going outdoors within 1 hour of scanning; even the temperature of the scanner room was regulated to obtain the most precise measurements (Thrippleton et al., 2014). These methods may be the reason for low error in repeated measurements (0.14°C), with less deviation between successive measurements. Of course, not all studies have the capacity to limit such phenomena, and one could argue that varied reference data in both sexes and across a varied number of experimental situations is imperative for fully understanding MRSI-t-based temperature estimates. Additionally, the calibrated equation we used for deriving T_CRE_ was developed on a more dated scanner model than that used for our study. There is disagreement over whether a calibrated formula derived phantom data is a worthwhile endeavor (Verius et al., 2019; Annink et al., 2020). The difference is minimal for short TE MRSI-t, but significant for long TE MRSI-t, resulting in a mean difference between derivations of up to 0.15°C (Annink et al., 2020). Given this small but possibly significant difference, future studies would benefit from developing a calibrated temperature formula that accounts for conditions at their particular scanner. It is worth noting that a calibration would only impact the intercept of the temperature calculation, with no impact on the slope. Thus, calibration would have little impact on outcomes if all participants’ data are acquired with the same sequence and identical temperature equation.

### 4.3 Conclusions

MRSI-t is a reliable and reproducible approach to measuring brain temperature, though future studies of larger sample size with more repeated acquisitions over long duration are necessary. We must also determine whether MRSI-t measurements of brain temperature are sensitive to the phenomenon we are attempting to visualize. Since this study included healthy participants only, the relationship between brain temperature and neuroinflammatory metabolites could not be fully assessed. Thus, the reference data this study provides must be applied to assessing patients with focal neuroinflammation. Before MRSI-t-based temperature can be utilized clinically, it is imperative to determine 1) if this tool can isolate focal brain temperature increases in regions of neuroinflammation and 2) if it can differentiate those with neuroinflammatory pathophysiology from those who are healthy.

## Data Availability

De-identified data will be made available upon reasonable request with IRB and data sharing approvals in place.

## 6 ETHICS STATEMENT

All study procedures were approved by the UAB Institutional Review Board. Written informed consent was obtained from all participants before initiating the protocol.

## 8 AUTHOR CONTRIBUTIONS

All authors listed have made substantial, direct, and intellectual contributions to the work. AAS, RN, and JPS: methodology and writing – review and editing. AAS and RN: data acquisition and processing. AAS and AM: methodology, data curation, and data visualization. CM, AM, JY: review and editing. AAS: formal analysis, writing—original draft. JPS: project supervision, funding acquisition, conceptualization, review, and writing—review and editing.

## 10 ACKNOWLEDGEMENTS

The authors thank Dr. Adam Goodman for helpful discussions regarding data analyses and visualization. This study was presented in part at the Annual Meeting of the Organization for Human Brain Mapping, Montreal, CA, 2020 (virtual presentation).

